# Development and Evaluation of Machine Learning Models for the Detection of Emergency Department Patients with Opioid Misuse from Clinical Notes

**DOI:** 10.1101/2024.12.11.24318875

**Authors:** Usman Shahid, Natalie Parde, Dale L. Smith, Grayson Dickinson, Joseph Bianco, Dillon Thorpe, Madhav Hota, Majid Afshar, Niranjan S. Karnik, Neeraj Chhabra

## Abstract

**Objectives:** The accurate identification of Emergency Department (ED) encounters involving opioid misuse is critical for health services, research, and surveillance. We sought to develop natural language processing (NLP)-based models for the detection of ED encounters involving opioid misuse.

**Methods:** A sample of ED encounters enriched for opioid misuse was manually annotated and clinical notes extracted. We evaluated classic machine learning (ML) methods, fine-tuning of publicly available pretrained language models, and a previously developed convolutional neural network opioid classifier for use on hospitalized patients (SMART-AI). Performance was compared to ICD-10-CM codes. Both raw text and text transformed to the United Medical Language System were evaluated. Face validity was evaluated by term feature importance.

**Results:** There were 1123 encounters used for training, validation, and testing. Of the classic ML methods, XGBoost had the highest AU_PRC (0.936), accuracy (0.887), and F1 score (0.863) which outperformed ICD-10-CM codes [accuracy 0.870; F1 0.830]. Logistic regression, support vector machine, and XGBoost models had higher AU_PRC using transformed text, while decision trees performed better using raw text. Excluding XGBoost, fine-tuned pre-trained language models outperformed classic ML methods. The best performing model was the fine-tuned SMART-AI based model with domain adaptation [AU_PRC 0.948; accuracy 0.882; F1 0.851]. Explainability analyses showed the most predictive terms were ‘heroin’, ‘opioids’, ‘alcoholic intoxication, chronic’, ‘cocaine’, ‘opiates’, and ‘suboxone’.

**Conclusions:** NLP-based models outperform entry of ICD-10-CM diagnosis codes for the detection of ED encounters with opioid misuse. Fine tuning with domain adaptation for pre-trained language models resulted in improved performance.

## Introduction

Drug overdose is the leading cause of accidental death in the United States with the majority involving an opioid.^1^ A critical healthcare setting for the initiation of treatments and medications for opioid use disorder (OUD) is the Emergency Department (ED).^2^ While treatments exist to decrease mortality related to opioid use, only a minority of patients with OUD are engaged in medical treatment.^3,4^ As people with OUD disproportionately utilize emergency services, ED encounters serve as valuable opportunities for interventions and linkage to outpatient healthcare resources. The accurate identification of patients at risk for OUD in the ED setting is critical to providing these life-saving treatments.

Current methods to identify patients with opioid misuse, defined as taking an opioid in a manner other than prescribed or using illicit opioids, rely on clinical interactions and documentation, often in the form of diagnosis codes.^5–8^ Universal manual screening for opioid misuse in the ED has been proposed but is highly resource-intensive and infrequently performed.^5,9^ Prior work has shown that documentation of opioid misuse by International Classification of Diseases, 10^th^ revision, Clinical Modification (ICD-10-CM-CM) diagnosis code is highly insensitive and fails in identifying a large proportion of patients that would benefit from interventions.^10^ The low sensitivity of ICD-10-CM based approaches for patient identification has impacts on research, surveillance, resource allocation, and clinical services in the healthcare setting. Methods in natural language processing (NLP) and machine learning (ML) to process clinical notes for clinical workflows have shown promise to build tools for screening opioid misuse in the inpatient setting.^11–13^ Such models rely on inpatient hospital documentation from the electronic health record (EHR), which far exceeds the amount of documentation performed during a typical ED encounter. It is currently unknown if NLP models trained using machine learning can be successfully domain adapted for screening opioid misuse in the ED setting.

The goal of the current study is to develop and evaluate NLP-based machine learning models to screen patients for opioid misuse during their ED encounter. We hypothesized that these models would outperform diagnosis codes in the identification of patients with opioid misuse, potentially highlighting the utility of such models for health services delivery, research, and surveillance.

## Methods

### Setting and Cohort Development

University of Illinois Hospital and Health Sciences System (UIHealth) is comprised of a 462-bed tertiary referral hospital with a 30-bed emergency department providing care across 45,000 patient encounters annually. It is located on the westside of Chicago, Illinois within an urban area that has among the highest opioid-related mortality in the United States.^14,15^ UIHealth has maintained Epic as its EHR vender since 2020 with a clinical data warehouse (CDW) of all patient encounters and associated data. The retrospective observational cohort of ED encounters used for training and testing of classifier models was drawn from the notes and reports extracted from the CDW with inclusion criteria of age greater than or equal to 18 years and encounter origination in the ED. A train/validation/test corpus was developed using a sample of approximately 1200 ED encounters. The sample was enriched for suspected opioid misuse using previously described methods involving sampling ED encounters with a positive urine opiate screen without coexisting opioid prescription or medication administration on hospital intake or an opioid-related ICD-10-CM diagnostic code and matching with encounters lacking these criteria.^10,16^ Enrichment was pursued to allow a balanced dataset for more efficient model training and improved classification. Encounters matching the criteria during the study period of September 2020 to March 2023 were identified and matched in a 1:1 ratio on disposition status (hospital admission or discharge) with encounters lacking previously described criteria or additional criteria placing the patient at risk for opioid misuse: an ICD-10- CM code for a chronic pain diagnosis, an order for naloxone, or an order for a urine drug regardless of result. We matched on disposition in lieu of age and sex to minimize bias from increased documentation, testing, and entry of ICD-10-CM diagnosis codes based on admission status and because age and sex are potentially important predictor variables for opioid misuse.

### Manual Annotation

The dataset was further processed with expert manual annotations. As opioid misuse represents a heterogeneous pattern of drug use and is difficult to categorize solely from variables within the EHR, potential cases and non-cases were manually annotated in a blinded format using a structured manual annotation schema. For the purposes of annotation, opioid misuse was defined as taking an opioid for reasons other than prescribed or as an illicit drug, consistent with definitions from the National Institute of Drug Abuse (NIDA) and the National Survey on Drug Use and Health (NSDUH).^6–8^ Annotators underwent a one-hour online training session with an expert in emergency addictions care (NC). They were required to achieve an interrater reliability kappa of greater than 0.80 with the expert prior to independent annotation. Annotators again completed cases in parallel with the expert and were given additional cases to annotate only after achieving threshold interrater reliability following every 100 cases independently annotated. Case details were extracted from the EHR by the annotators into a structured REDCap data collection form for each encounter.^17,18^

Using previously described methods, the presence of opioid misuse was determined using a 5- point Likert scale indicating the probability of opioid misuse which included the categories of definite, highly probable, probable, definitely not, and uncertain.^10,11^ Probable cases required either: 1) history of opioid misuse evident in clinical notes but no current documentation for the encounter; 2) provider mention of drug-seeking behavior; or 3) evidence of other drug misuse, except alcohol, in addition to prescription opioid use. Highly probable cases had either more than one probable case criteria or provider mention of suspicion of opioid misuse. Definite cases were classified by either patient self-report of opioid misuse or documentation by provider of patient misusing an opioid. Cases lacking any of the above criteria were classified as ‘definitely not.’ For classification, cases annotated as probable, highly probable, or definite were categorized as exhibiting opioid misuse.

### Model Development

Encounters comprising the cohort were sorted chronologically by encounter date and divided into training, validation, and testing sets at a ratio of 70/15/15. We evaluated multiple model architectures, all of which can be broadly categorized as either classic (feature-based) machine learning and neural methods. Classic machine learning methods included logistic regression, support vector machines, decision trees, and eXtreme Gradient Boosted (XGBoost) trees. Neural methods utilized publicly available pre-trained language models or a previously trained convolutional neural network opioid classifier for use on hospitalized patients (SMART-AI).^13^ We experimented with two variants of the SMART-AI classifier. Firstly, we used the classifier as a feature extractor and trained an MLP classifier on these features. In the second variant, we fine-tuned this model end-to-end on our dataset to improve domain adaptation.^19^ The pre-trained language models included Bidirectional Encoder Representations from Transformers (BERT), BioBERT, Longformer, Clinical Longformer, and Clinical BigBird. BERT is an encoder-only transformer model developed by Google and Longformer is a modified transformer model which can account for longer text sequences for use on general text.^20,21^ BioBERT, Clinical Longformer, and Clinical BigBird are domain-adapted language models pre-trained on medical corpora.^22,23^. For neural models, we performed domain adaptation with fine-tuning of the pre-trained model on the training data by using the averaged embeddings from the final hidden layer of the pre-trained model and added a Multi-Layer Perceptron classifier on top with one hidden layer of size 256 and dropout layers with probability 0.3. Rectified Linear Unit (ReLU) was used as the activation function between intermediate layers and final probabilities were obtained using the sigmoid function, similar to the binary output provided by a logistic regression classifier.

### Text Processing

All clinical documents originating from individual ED encounters were concatenated in chronological order, including notes from staff with direct interaction with patients in the ED such as physicians and nurses. For each machine learning method, we evaluated both the use of raw text as well as transformed text (feature engineering), where applicable. For classic machine learning methods, text went through minimal preprocessing by converting to lower case and breaking the text into unigram (single word, top 10000 unigrams by frequency) feature vectors to represent individual variables. No feature engineering was performed for fine-tuning of pre-trained language models and the natural language of the text was used up to the context length of the language model (e.g., 512 for BERT based and 4096 for Longformer based models)

In addition to the raw text features, the text was also mapped to medical concepts and converted into structured codes, referred to as concept unique identifiers (CUIs). The text was mapped to the National Library of Medicine’s Unified Medical Language System (UMLS) concept unique identifier (CUI) codes using the clinical Text And Knowledge Extraction System (cTAKES). cTAKES is an open-source NLP engine which identifies named entities in raw text and maps them to the UMLS.^24,25^ This transformation accounts for negation and outputs CUI codes. An additional feature of cTAKES transformation is the stripping of protected health information from the raw text as part of processing into named entities and CUIs as well as bringing together semantically similar terms to the same medical concept using the UMLS Metathesaurus (i.e, opioid use disorder and OUD). We chose to evaluate both raw and transformed text. This is because raw text retains the contextual information in a sentence, while the transformed text using cTAKES maps similar terms to a single CUI thus negating some variability due to individual clinician vocabulary and word choice.

### Statistical Analysis

Models were compared along multiple metrics with the primary being Area Under the Precision Recall Curve (AU_PRC). The AU_PRC considers both precision (positive predictive value) and recall (sensitivity) and was chosen as the primary metric given the importance of identifying all positive cases and better accounting for imbalanced datasets. We also considered other metrics such as accuracy, F1 score, area under the receiver operator curve (AU_ROC), and language model complexity as determined by the number of parameters. We evaluated the number of parameters (i.e., model weights) for language models since a higher number of parameters can represent a need for more significant computational resources. We selected our final model as the one with the most favorable metrics, prioritizing AU_PRC. In the case of similarly performing models, parsimony was prioritized. We also compared model performance against clinical detection of opioid misuse ICD-10-CM diagnosis codes.^26,27^ Entry of ICD-10-CM codes represent clinician identification of opioid misuse. ICD-10-CM codes are commonly used for problem list generation, billing, surveillance, and research. They represent a baseline that any potential classification model should outperform. As a binary variable, we only determined a subset of metrics for ICD-10-CM code performance: precision, recall, F1 score, and accuracy.

### Feature/Variable Importance

To evaluate face validity of the final model, we estimated feature importance for the best-performing model on the held-out test set. Given the black box nature of many neural methods, we used Local Interpretable Model-agnostic Explanations (LIME) to evaluate the top 25 features which were most predictive of opioid misuse.^28^ Designed for explainable artificial intelligence, the LIME algorithm fits multiple surrogate linear regression models to approximate a machine learning model and evaluates feature importance for each prediction in the form of beta coefficients, referred to as LIME scores. We used LIME to evaluate the features which predicted whether a patient was positive or negative for opioid misuse for each observation in the held-out test set.

All analyses were performed in Python (version 3.12) using PyTorch (version 2.4).^29^ This study was reviewed and exempted from review as non-human subjects research by the institutional review board of the primary institution. The study conforms, where appropriate, to Transparent Reporting of a multivariable prediction model for Individual Prognosis Or Diagnosis + Artificial Intelligence (TRIPOD+AI) guidelines (Appendix A).^30^

## Results

Of the initial 1200 annotated ED encounters comprising the study cohort, 77 encounters were removed for enhanced privacy protections associated with patient chart (62) or lack of any text documentation (15), leaving 1123 encounters for training, validation, and testing. Of the total 1123 cases, 570 were labeled as opioid misuse positive and 553 labeled negative. Demographic and other information for the cohort is shown in Table 1. Encounters were divided temporally into training, validation, and testing sets with 786, 168, and 169 encounters, respectively. In the test set, there were 75 encounters labeled positive for opioid misuse and 94 labeled negative.

**Table 1.** Demographics of Cohort.

| Variable | n (%) |
| --- | --- |
| Age |  |
| 18-34 | 285 (25.38%) |
| 35-49 | 255 (22.71%) |
| 50-64 | 360 (32.06%) |
| 65+ | 223 (19.86%) |
| Sex |  |
| Female | 543 (48.35%) |
| Male | 580 (51.65%) |
| Race/Ethnicity |  |
| Non-Hispanic Black | 636 (56.63%) |
| Hispanic or Latino | 294 (26.18%) |
| Non-Hispanic White | 159 (14.16%) |
| Other | 25 (2.23%) |
| Patient Declined | 9 (0.80%) |
| Insurance |  |
| Medicaid | 665 (59.22%) |
| Medicare | 245 (21.82%) |
| Private | 130 (11.58%) |
| Other | 18 (1.60%) |
| No Information | 65 (5.79%) |
| ED Disposition |  |
| AMA | 77 (6.9%) |
| Admit | 487 (43.4%) |
| Discharge | 417 (37.13%) |
| Expired | 2 (0.18%) |
| Observation | 59 (5.25%) |
| Sent to operating room | 2 (0.18%) |
| Other | 79 (7.0%) |

Notes: ED, Emergency Department; LWBS, left without being seen by medical provider; AMA, left against medical advice, eloped, or left without being seen by provider. “Other” disposition category includes transfers to other facilities, transfers to behavioral health, transfers to obstetrics, and unspecified dispositions.

Of the classic machine learning methods, XGBoost had the highest AU-PRC (0.936), accuracy (0.8876), and F1 score (0.8633) on the held-out test set (Table 2). Logistic regression, support vector machine, and XGBoost-based models all had higher AU_PRC when used with CUIs, while only decision tree models had improved metrics with untransformed text. Of the classic machine learning methods, only XGBoost models outperformed ICD-10-CM codes in recall, F1 score, and accuracy.

**Table 2:**
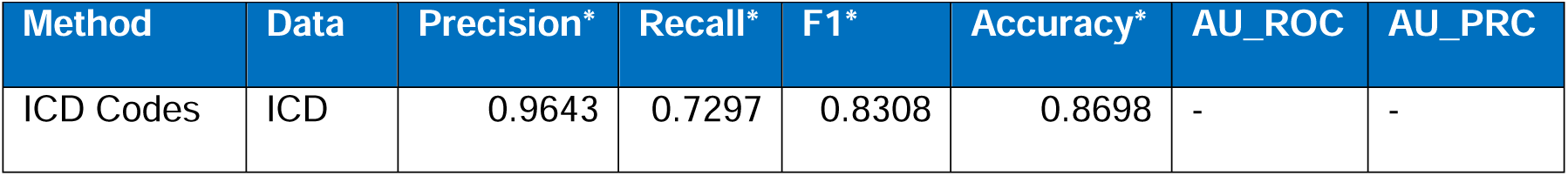
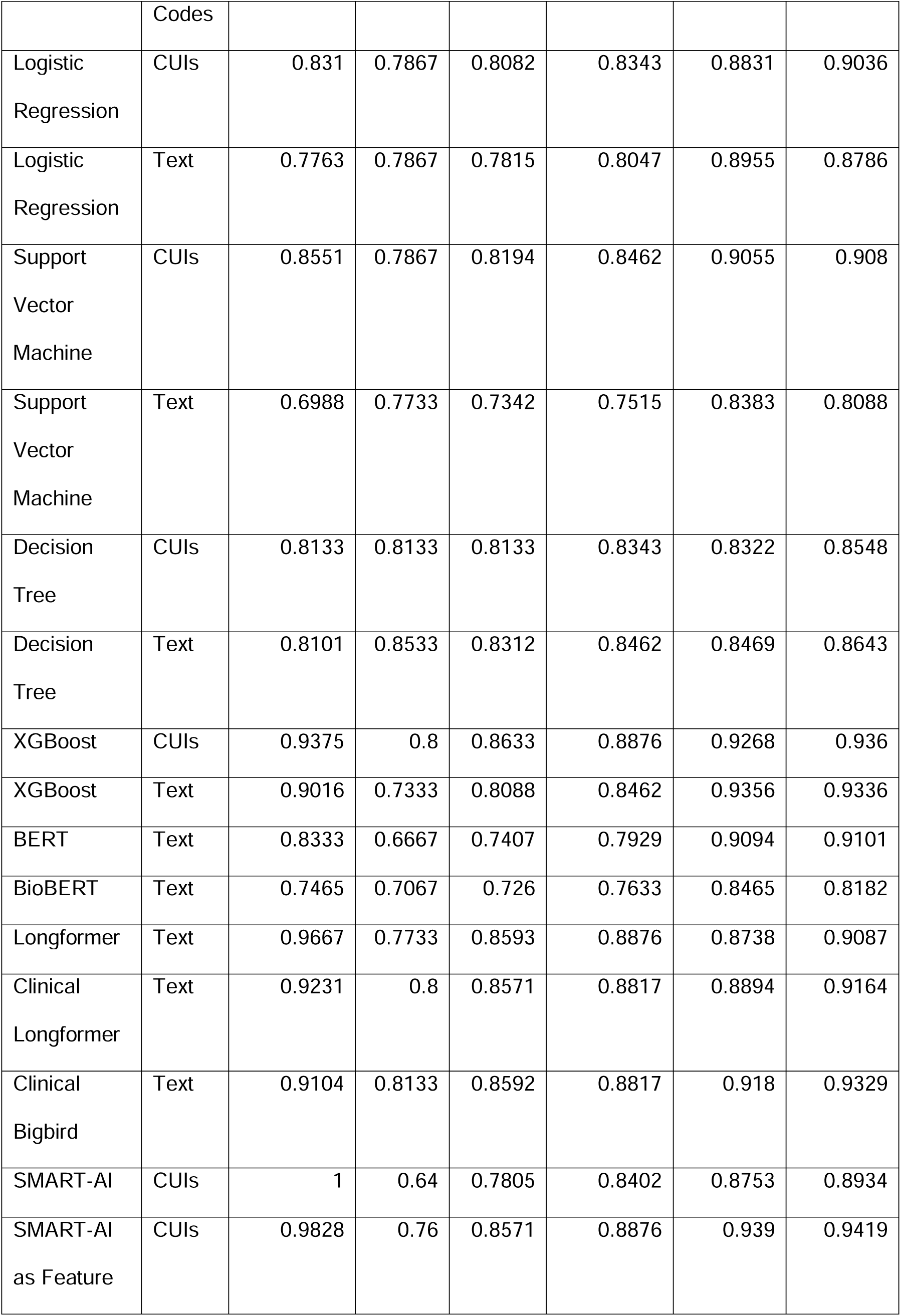
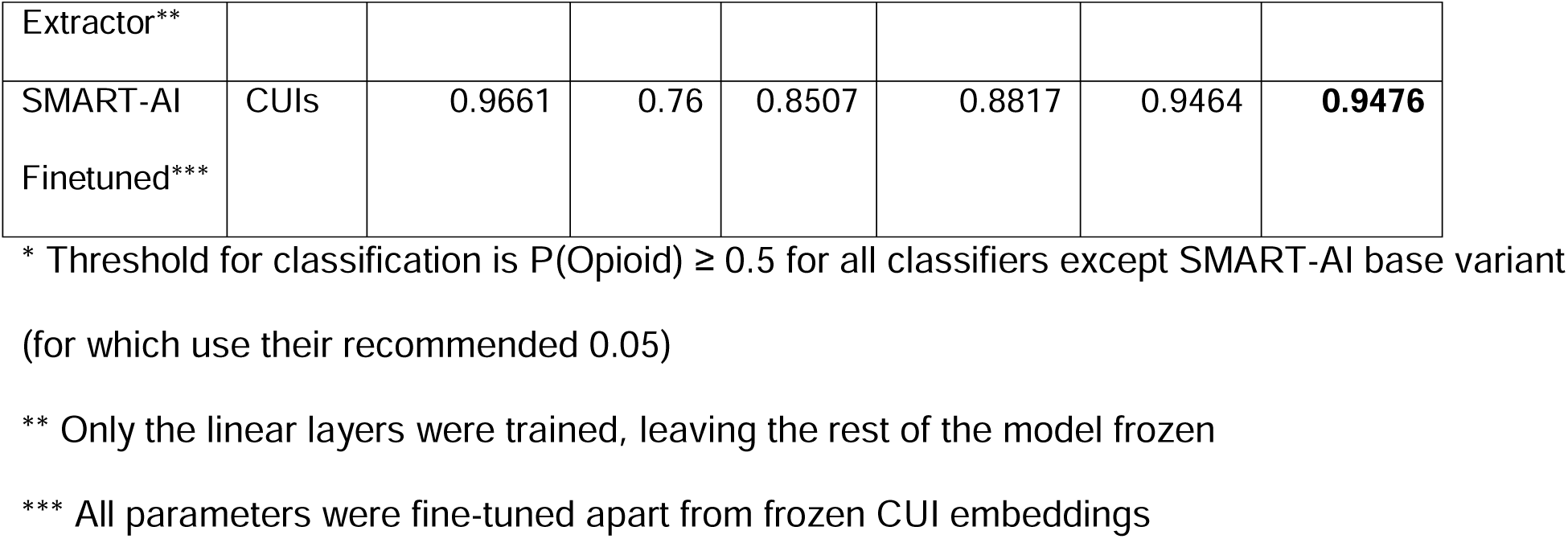
results of classifiers by method and data preparation.

With the exception of XGBoost, the neural classifiers utilizing pre-trained language models generally outperformed the classic machine learning methods in AU_PRC. The best performing models were the SMART-AI based models which used CUIs as inputs. The SMART-AI model that underwent transfer learning with domain adaptation through fine tuning with frozen CUI embeddings had the highest AU_PRC and AU_ROC of all models evaluated.

We considered model complexity as well, and we present those findings in Table 3. The SMART-AI based models had fewer parameters than all pretrained language models. Explainability analyses using LIME show the CUIs more predictive of opioid misuse were ‘heroin’, ‘opioids’, ‘alcoholic intoxication, chronic’, ‘cocaine’, ‘opiates’, and ‘suboxone’ (Figure 1).

**Figure 1:**
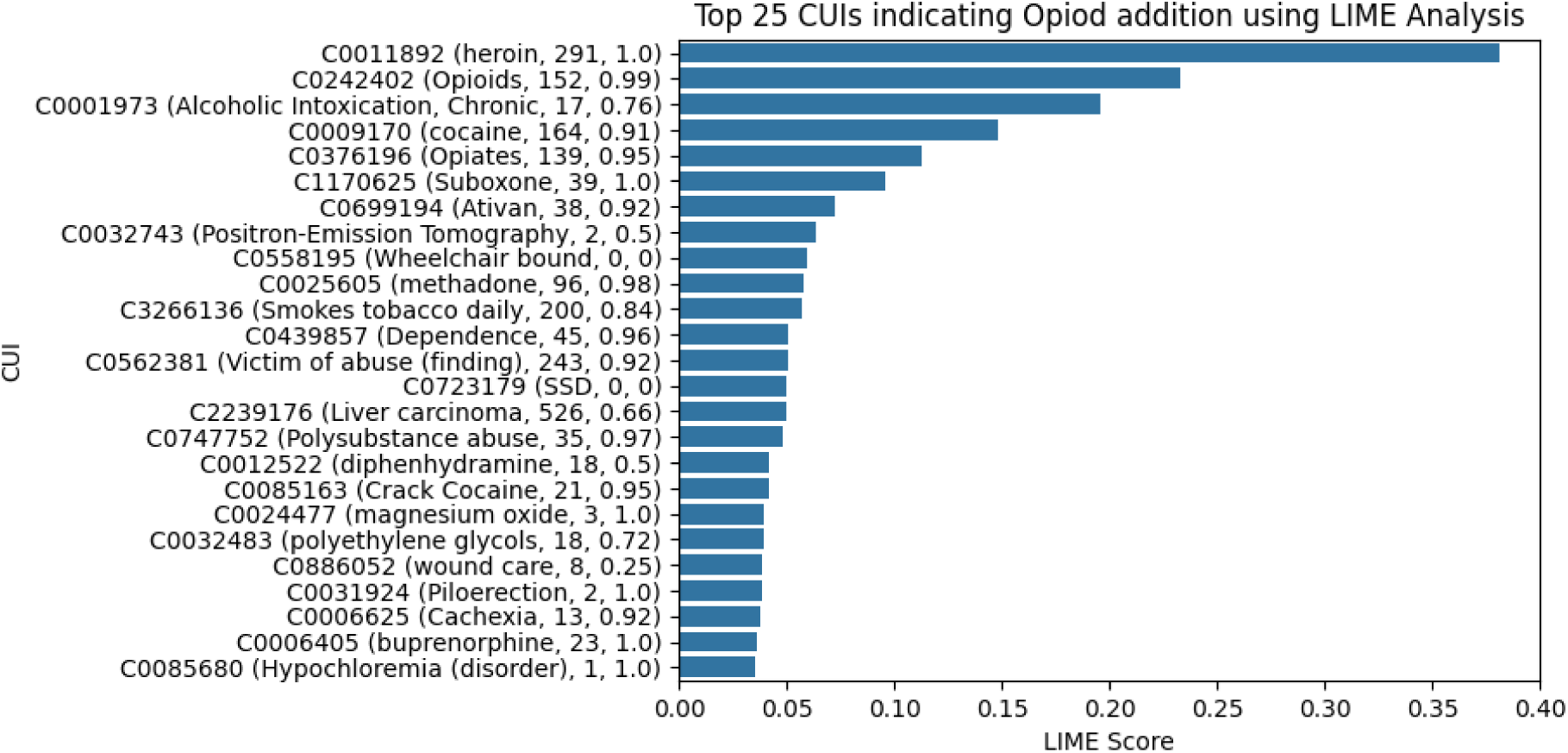
Feature Importance for predicting opioid misuse *Descriptions along the y-axis are of the following format: concept unique identifier (text descriptions from the National Library of Medicine’s Unified Medical Language System, number of encounters from the training set with the concept unique identifier, proportion of encounters with the concept unique identifier that was labeled as displaying opioid misuse). LIME scores range from 0-1 with higher scores indicating higher predictive power on the test set.

**Table 3:** Neural Classifier Parameters.

| Model | Data | No. of Parameters |
| --- | --- | --- |
| BERT | Text | 109 M |
| BioBERT | Text | 108 M |
| Longformer | Text | 148 M |
| Clinical Longformer | Text | 148 M |
| Clinical Bigbird | Text | 127 M |
| SMART-AI | CUI | 12.15 M |
| SMART-AI as Feature Extractor | CUI | 12.4 M |
| SMART-AI Finetuned | CUI | 12.4 M |

## Discussion

In this evaluation of model-based detection of patient encounters with opioid misuse in the ED setting, the best performing NLP classifiers outperformed ICD-10-CM codes for the detection of opioid misuse. The best performance was noted from the models which were trained for the detection of opioid misuse in hospitalized patients and adapted to this domain by employing transfer learning techniques. These models were also more lightweight, based on the number of parameters, than those using pretrained language models. In general, models based on text transformed to a standard UMLS lexicon performed as well or better than those using raw text in comparable settings. Some of the pretrained language models, especially Clinical Bigbird, had comparable performance, most likely due to its large context window and pretraining on medical text (MIMIC-III clinical notes).^23^ It is important to note that XGBoost, a lightweight classifier, stood out among classical machine learning methods by delivering comparable performance to neural approaches without necessarily requiring text pre-processing with cTAKES.

Model-based detection of patients with opioid misuse has implications in healthcare delivery, disease surveillance, and research. The accurate detection of patients likely to benefit from opioid-specific interventions is critical in the ED setting. Universal manual screening is infrequently performed due to being resource intensive and because it may not be ideally suited for the emergency setting where there are multiple competing clinical concerns in a highly time-sensitive environment.^9^ Diagnosis code entry, which serves as a surrogate marker for the detection of opioid misuse, is highly insensitive.^10,31,32^ Current methods of patient detection would leave many patients that would likely benefit from medical treatment for opioid use disorder without being detected. As medical treatment is already uncommon among patients with opioid misuse in the ED, improved detection is critical.^33^ Similarly, surveillance for opioid misuse among ED patients has implications for resource allocation and programmatic funding. Underdetection leaves already resource-strapped environments with a diminished ability to advocate for expanded resources necessary to care for patients with opioid use disorder. As ICD-10-CM codes are highly relied upon for cohort development in research, the improved detection with NLP-based models would allow for decreased selection bias and improved validity of research reliant on diagnosis codes for cohort discovery.

In the current study, we evaluated both the use of raw text and transformed text as features for candidate models. Both have theoretical benefits and drawbacks. Raw text, when used with pre-trained transformer-based models, has the benefit of context from surrounding words to improve classification. It also avoids an additional layer of computational burden that occurs with text pre-processing. In our analysis, we evaluated transformed text to a standardized lexicon from UMLS using cTAKES. Although context is lost by using cTAKES as it transforms named entities without regard for nearby terms, advantages of using cTAKES are that it has the ability to: 1) map disparate terminology to single distinct concepts; and 2) remove identifying protected health information (PHI) such as names and dates. The benefits of mapping to CUIs negate some of the effects of individual variations in vocabulary, allowing similar concepts to be aggregated. The removal of PHI carries implications for model deployment as multiple institutions can theoretically share lists of deidentified CUIs from clinical encounters for model processing, allowing for the accurate surveillance for disease processes by public health entities across multiple institutions without compromising patient privacy.

Among similarly performing models, the simpler model is preferred due to explainability, scalability, and transportability. Pretrained language models, while computationally expensive, did not outperform the SMART-AI classifier, which has fewer parameters by an order of magnitude. In addition, the baseline SMART-AI classifier without adaptations was outperformed by a version fine-tuned on ED-specific data, which speaks to the successful adaptation to this domain using transfer learning from external sources. Note that most of the SMART-AI parameters (approximately 11.2M) were pre-trained CUI embeddings. Evaluation of black-box neural network-based clinical models for interpretability is an important consideration prior to clinical deployment. The domain-adapted SMART-AI model underwent evaluation for interpretability with the most important CUIs in predicting opioid misuse representing concepts with high face validity. Some terms with decreased association with opioid misuse, such as ‘Positron Emission Tomography,’ are likely related to features noted in sparse training data for either the original or adapted classifier or a result of term overlap where some strings may be mapped to incorrect concepts. The next steps before clinical use of a domain-adapted SMART-AI model involve subgroup validation and recalibration on a larger cohort of ED encounters.

All experimental results should be interpreted within the context of their limitations. As ICD-10- CM codes were a portion of the criteria for cohort development, performance metrics for ICD- 10-CM codes were likely inflated. For experiments with pretrained language models, we were limited by the context window limitations of the individual models. Preprocessing of text with cTAKES requires a license and carries the potential limitation of mapping of named entities to incorrect CUIs. All models designed for clinical use should undergo bias and fairness assessments to ensure equity. Bias assessments for the current study were unrevealing owing to the small test set size and should be evaluated on a larger sample with appropriate mitigation techniques prior to clinical deployment.

## Conclusion

Natural language processing-based models can outperform entry of ICD-10-CM diagnosis codes in the detection of encounters with opioid misuse in the ED setting. Fine tuning with domain adaptation for pre-trained language models resulted in improved performance of our opioid misuse classifier.

## Data Availability

All data produced in the present study are available upon reasonable request to the authors subject to patient privacy protections

